# Retrospective feasibility study of *Mycobacterium tuberculosis* modified lipoprotein as a potential biomarker for TB detection in children

**DOI:** 10.1101/2021.03.26.21254411

**Authors:** Xinxin Yang, Matthew F. Wipperman, Sharon Nachman, Nicole S. Sampson

**Affiliations:** Department of Chemistry, Stony Brook University, Stony Brook, NY 11794-3400; Chronus Pharmaceuticals, Inc., 25 Health Sciences Drive, Stony Brook, NY 11790; Clinical and Translational Science Center, Weill Cornell Medicine, New York, NY 10065; Department of Pediatrics, Renaissance School of Medicine, Stony Brook University, Stony Brook, NY 11794-3400

**Keywords:** Childhood TB, HIV co-infection, diagnosis, sandwich ELISA, lipoprotein, low-density lipoprotein, apoB, biomarker

## Abstract

**Background:** Current TB diagnostic methods available have been developed for adults and development efforts have neglected the differences in disease and sampling that occur between adults and children. Diagnostic challenges are even greater in HIV co-infected children and infants.

**Methods and results:** We established a sandwich ELISA assay to detect *Mycobacterium tuberculosis* modified lipoprotein (TLP) ex vivo in plasma. The study population contains plasma samples from 21 patients with active TB and 24 control samples with no TB, collected in the International Maternal Pediatric Adolescent AIDS Clinical Trails (IMPAACT) P1041 study. Retrospective analysis was performed and the result demonstrate that TLP level is associated with TB disease.

**Conclusions:** Plasma levels of TLP associate with active TB disease in HIV positive subjects and can be used as an indicator for TB detection in children.

## BACKGROUND

Childhood TB is estimated to contribute 12% of the disease burden, with approximately 10 million cases in 2019 worldwide [1]. Existing gold-standard diagnostic culture tools fail to confirm TB in most children, who typically have low bacterial counts and cannot produce sputum like adults. Diagnostic challenges are great in HIV co-infected children and infants [2], where the clinical presentation of pulmonary TB may be non-specific [3], acute [4, 5], and chest radiographs may be atypical [2]. Although rates of bacteriological confirmation appear similar in HIV-infected and uninfected children [6], diagnostic delay and HIV-related immune pathology contribute to higher rates of TB disease progression with increased disease severity, morbidity, and mortality [7-10]. Infants, young children, and HIV-infected children are at increased risk of developing TB following infection and of disseminated or severe disease, including TB meningitis [11]. Because TB in children is typically paucibacillary, even if a sputum sample or gastric aspirate is obtained, the utility of PCR based diagnostic methods like GeneXpert is limited. More sensitive and child-friendly diagnostic tools are urgently needed to diagnose TB in children [12].

*Mycobacterium tuberculosis* (*Mtb*) is an intracellular pathogen that is capable of surviving and replicating within macrophages, the frontline of the innate host defense, and has evolved multiple mechanisms to escape these immune cells [13]. One of the immune evasion strategies of *Mtb* is the deregulation of lipid metabolism, leading to the formation of foamy macrophages (FM), a hallmark of granulomata in tuberculosis lesions [14]. However, only pathogenic mycobacterial strains including *M. tuberculosis* [15], *M. avium, M. abscessus, or M. bovis*, but not non-pathogenic mycobacterial strains, like *M. smegmatis*, induce foam cell formation upon infection [16, 17]. Foam cells are associated with chronic inflammation in many metabolic diseases and certain cancers besides infectious diseases [18, 19]. For example, foamy macrophages are critical to the initial formation, development, and instability of atherosclerotic plaques and are therefore therapeutic targets in atherosclerosis. Foam cell formation is induced primarily by malondialdehyde-modified low density lipoprotein (MDA-LDL) and not by native or extensively oxidized LDL in atherosclerosis [20]. Elevated plasma levels of MDA-LDL in patients are associated with acute coronary syndromes and are used as diagnostic tools clinically [21].

In this study we identified human lipoprotein modification specific to exposure to *Mtb*. The pathogen modified host lipoprotein, TLP, is detectable ex vivo in plasma using a sandwich ELISA assay. We further evaluated the association between the presence of TLP and TB disease status in children.

## METHODS

### Preparation of malondialdehyde-conjugated LDL (MDA-LDL) [20] and Mtb-modified lipoprotein (TLP)

HepG2 human liver cells (ATCC HB-8065) were grown to 80% confluence in HepG2 growth media (DMEM, 10% fetal bovine serum, 20 mM L– glutamine, 100 U/mL penicillin, 100 μg/mL streptomycin, and 10 mM HEPES). Cells were grown for 4-5 days and culture supernatants were then harvested and concentrated. LDL particles were separated by density gradient ultracentrifugation and desalted by ultrafiltration through a 100-kDa molecular weight filter. MDA-LDL was prepared by incubation 2 mg protein/mL of LDL with 2 μM acrolein at 37 °C for 24 h under nitrogen atmosphere. *Mycobacterium tuberculosis* was cultured in Middlebrook 7H9 (broth) supplemented with 0.2% glycerol, 0.5% BSA, 0.08% NaCl, 0.05% (v/v) tyloxapol to OD∼0.7. LDL was added and the culture was incubated at 37 °C for 7 days. TLP was isolated from the culture supernatant, concentrated and washed with PBS.

### Agarose gel electrophoresis and western blot analysis

Agarose gel electrophoresis was performed in 0.06 M barbital buffer (pH 8.6). The gel was 0.8% and stained with Sudan Red 7B. Lipoproteins were separated on a 6% sodium dodecyl sulfate polyacrylamide gel electrophoresis (SDS-PAGE), proteins were transferred to a PVDF membrane and immunoblotted with specific antibodies. The primary antibodies used for western blot were monoclonal anti-LDL (MDA oxidized) antibody (Abcam) and polyclonal anti-apoB Ab (H-300, Santa Cruz). Membranes were treated with anti-mouse, or anti-rabbit IgG HRP conjugates as secondary antibodies.

### THP-1 macrophage preparation and treatment

THP-1 cells (ATCC TIB-202) were maintained in RPMI-1640 medium containing heat-inactivated 10% fetal bovine serum (FBS), 0.05 mM 2-mercaptoethanol, 100 U/mL penicillin and 100 mg/mL streptomycin (RPMI complete medium). The cells were plated into 6-well plates containing coverslips (1.5×10^6^ /well), treated with phorbol 12-myristate 13-acetate (PMA, 150 nM) in RPMI complete media. Media was replaced with fresh RPMI complete media at 48 hours after plating. At 72 hours after media change, THP-1 macrophages were treated with LDL, MDA-LDL and TLP at 200 μg/mL in PBS for 24 hours. All cells were incubated in a humid atmosphere at 37°C with 95% air and 5% CO_2_.

### Lipid body staining and immunostaining

Following lipoprotein treatment, macrophages were washed with PBS and fixed with 4% paraformaldehyde at room temperature for 30 minutes. Cells were stained with Oil Red O solution for 20 minutes at room temperature. The slides were then counterstained with haematoxylin and observed under an inverted microscope (Zeiss Axiovert 200M). Percent foam cell formation was quantified by counting stained versus total cells in 10 fields.

### Generation of anti-TLP antibodies

[22, 23]. Monoclonal antibodies against TLP were identified utilizing Human Combinatorial Antibody Library (HuCAL, Bio-Rad AbD Serotec GmbH). The HuCAL phage display library was depleted of antibodies that recognize intact LDL. The depleted library was panned for three rounds of binding, elution, and amplification to isolate antibodies specific for TLP. Binding of anti-TLP antibodies to control antigens including BSA, HSA, N1-CD33-His6 and intact LDL, was checked by indirect ELISA assays, with anti-TLP antibodies as primary antibodies and an anti-Fab-AP conjugate (Bio-Rad) as a secondary antibody.

### Sandwich ELISA assay

Briefly, 96 well plates were coated overnight at 4°C with 5 μg/mL capture antibody (AbD28580, amino acid sequence shown in Table S2) in phosphate buffered saline (PBS). Plates were washed with PBS containing 0.05% Tween-20 (PBST) 3 times, blocked with 3% BSA in PBST for 1 hour at room temperature. Then serial dilutions of standards and test samples in HISPEC assay diluent (Bio-Rad) were loaded and allowed to react for 2 hours at room temperature. Plates were then washed 3 times with PBST and treated with 2 μg/mL of HRP conjugated detection antibody (AbD28582) in HISPEC assay diluent for 1 hour at room temperature. After washing 6 times with PBST, the plates were developed with QuantaBlu fluorogenic peroxidase substrate kit (Thermo Scientific) for 30 min at room temperature. Fluorescence was recorded (ex. 320 ± 25nm, em. 430 ± 35nm).

### Subjects and samples

Retrospective analysis was performed on a total of 45 plasma samples from International Maternal Pediatric Adolescent AIDS Clinical Trails (IMPAACT) P1041 [24, 25]. P1041 was a Phase II/ III, randomized, double-blind, placebo-controlled clinical trial to evaluate the efficacy of isoniazid prophylaxis on TB disease and latent *Mtb* infection free survival in HIV-infected and HIV-exposed, but uninfected infants up to 192 weeks of follow up [25, 26]. Samples include 24 with noTB and 21 with TBDIS, all HIV infected. Patients without *Mtb* infection were categorized as noTB; patients with a positive tuberculin skin test but lacking any clinical, radiographic or laboratory evidence of disease caused by *Mtb* were categorized as latent TB infection (LTBI); and patients presenting clinical, radiographic or laboratory evidence of disease caused by *Mtb* were categorized as TBDIS. Plasma samples were collected, frozen and thawed before use.

### Statistical Analysis

Categorical variables were compared using t-student test, whenever appropriate. Non-parametric tests (Mann-Whitney) were used for non-normally distributed variables. Assay accuracy, including 95% confidence intervals, was assessed using sensitivity, specificity, predictive values and area under the ROC in the TB and non-TB groups. Statistical calculations were performed with GraphPad Prism® Software.

## RESULTS

### Mtb modified lipoprotein (TLP) is different from MDA-LDL

We purified and treated native LDL with *Mycobacterium tuberculosis* in vitro to obtain *Mt*b modified lipoprotein (TLP). Decreased mobility of TLP in agarose electrophoresis indicates that TLP is more positively charged than native LDL (Figure 1A). In contrast, MDA-LDL, one of the end products of lipid peroxidation, is more negatively charged than native LDL [20]. The increased size of TLP compared to native LDL was also confirmed by dynamic light scattering analysis (Table S1). Furthermore, TLP is recognized by anti-apolipoprotein B (apoB) antibodies but not anti-MDA-LDL antibodies, indicating TLP does not contain a malondialdehyde-modified apoB derivative (Figure 1B). We demonstrate that LDL is altered in the presence of *Mtb*, and the resulting TLP is distinct from the typical atherosclerotic species malondialdehyde-modified LDL (MDA-LDL) present in human blood [27].

**Figure 1.**
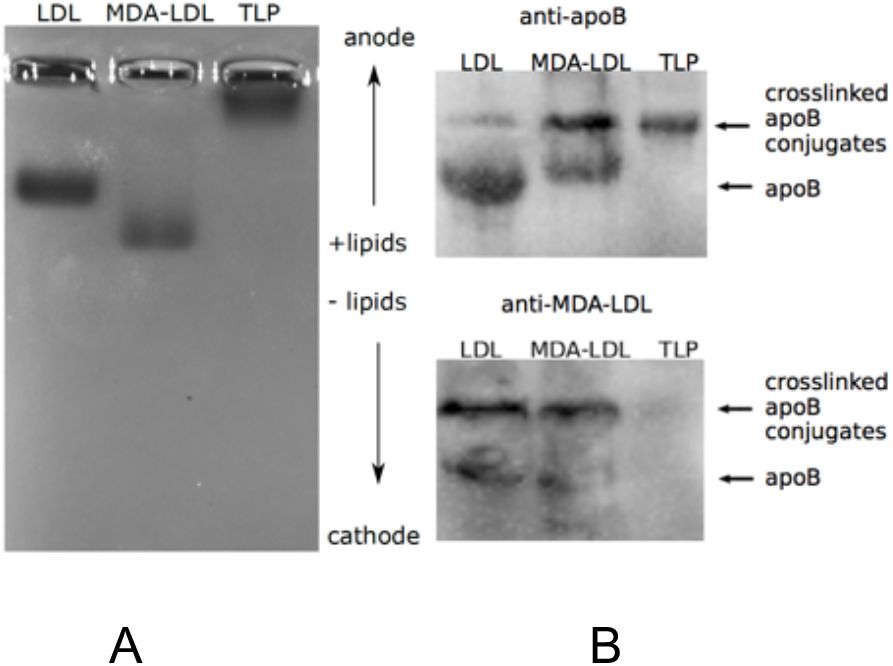
TLP is different from MDA-LDL. (A) an agarose gel electrophoresis showing TLP is larger in size and more positively charged than native LDL. (B) Immunoblots showing TLP is recognized by anti-apolipoprotein B (apoB) antibodies (top) but not anti-MDA-LDL antibodies (bottom). Images of the entire gel and blots are in Figure S1.

### Mtb modified LDL induces lipid body accumulation in macrophages

The induction of foamy macrophages have been reported as hallmarks in many pathologies associated with chronic proinflammatory stimuli including atherosclerosis [15]. It has also been reported that *Mtb* lipids induce the formation of giant multinuclear macrophages [28]. We therefore used foamy macrophage formation as a tool to assess the bioactive components that trigger a tissue response similar to granuloma formation in TB. Lipid bodies were stained with Oil red O. The buffer control showed no accumulation of lipid bodies and the cells remained rounded (Figure 2A). LDL treated macrophage showed some lipid accumulation, with a commensurate increase in cell size (Figure 2B). In contrast, MDA-LDL, commonly associated with atherosclerosis, treated macrophages exhibited more extensive accumulation of lipid bodies than the LDL treated macrophages. These macrophage phenotypes are consistent with previous reports of in vitro foamy macrophage formation [20, 29]. Most importantly, TLP treated macrophages had extensive lipid body accumulation per cell compared to macrophages treated with other LDLs (Figure 2 and S2B). Moreover, the TLP-treated macrophage morphology was distinct from other samples. The majority of the TLP-treated macrophages became multinucleated (Figure 2D and S2A) and developed long filopodia consistent with cell migration or exocytosis. Thus, we hypothesize that the modified LDL is detectable in vivo to serve as a biomarker for TB disease, and an antibody specific for recognition of TLP is an *Mtb*-specific indicator of infection and progression to (active) TB disease.

**Figure 2.**
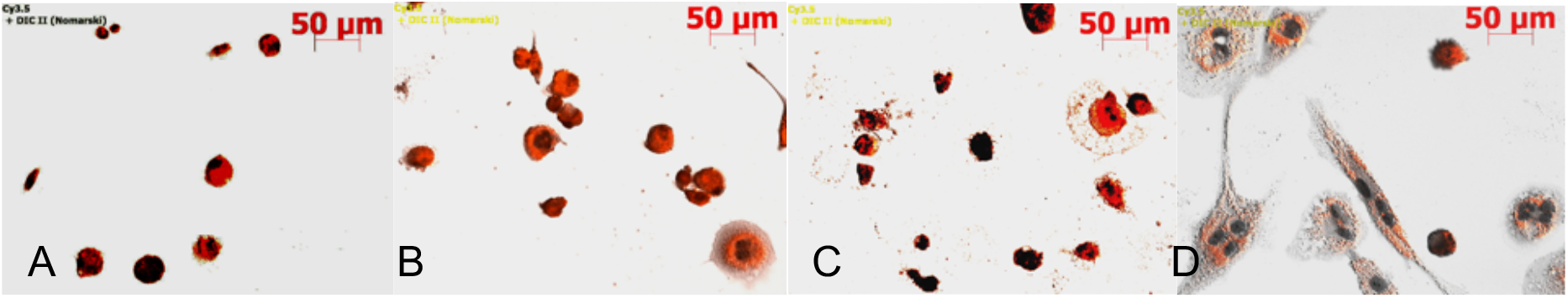
TLP induces lipid body accumulation in macrophages. Microscopic images of Oil Red O-hematoxylin stained THP-1 macrophage culture treated with PBS control (A), LDL (B), MDA-LDL (C) and TLP (D) at 200 μg/mL for 2 days.

### Anti-TLP monoclonal antibodies were generated and a TLP Sandwich ELISA assay was established

We generated 13 anti-TLP monoclonal antibodies (Fad-A-FH, bivalent Fab-bacterial alkaline phosphatase fusion antibody followed by FLAG^®^ and His6-tag) from the Human Combinatorial Antibody Library (HuCAL, Bio-Rad AbD Serotec GmbH) using in vitro selection and counter selection [22, 23]. HuCAL is a phage display library containing highly specific, fully human monoclonal antibodies, with DNA sequence encoded for each antibody fragment. The HuCAL phage display library was depleted of antibodies that recognize LDL (Figure S3). The depleted library was panned for three rounds of binding, elution, and amplification to isolate antibodies specific for TLP. Among the antibody hits selected, 13 antibodies were identified as unique by DNA sequencing. These antibodies were produced heterologously for further development. The cross-reactivity with native LDL, human serum albumin, bovine serum albumin, and His_6_ affinity tag was determined in an indirect ELISA format. Eight antibodies showed a specific signal for TLP at least 4-fold above native LDL background (Figure S3). We tested pairwise combinations of antibodies to identify antibody pairs that have non-overlapping epitopes suitable for use in a sandwich ELISA (Figure S4A). 5 sandwich pairs were identified with good signal over background and the assay conditions were optimized for the best pair, AbD28582 and AbD28580 (amino acid sequence shown in Table S2). We established the linear response of the sandwich ELISA (Figure S4B) to antigen spiked into pediatric plasma. TLP is specifically detected at >=4ppm in the presence of plasma LDL, which is around 10^6^ ng/mL [30].

### Levels of TLP are increased in patients with active TB compared to no TB controls

We assayed stored plasma samples from the HIV IMPAACT P1041 trial for the presence of TLP. The median plasma levels of TLP was 494 ng/mL in control subjects, and 2.7 fold higher in active TB subjects (Figure 3A). The difference between control subjects and active TB subjects is significant (p<0.001). The receiver operating characteristic (ROC) analysis describes the relationship between the sensitivity and specificity at any cut-off values. An ROC curve of TLP concentration for active TB versus control subjects is presented in Figure 3B. The area under the ROC curve (AUC) is 0.86 (95% CI=0.75-0.97, p<0.0001). An optimal cutoff of 1064 ng/mL rendered 71% sensitivity and 88% specificity. Alternatively, a cutoff of 1230 ng/mL provided 57% sensitivity and of 96% specificity.

**Figure 3.**
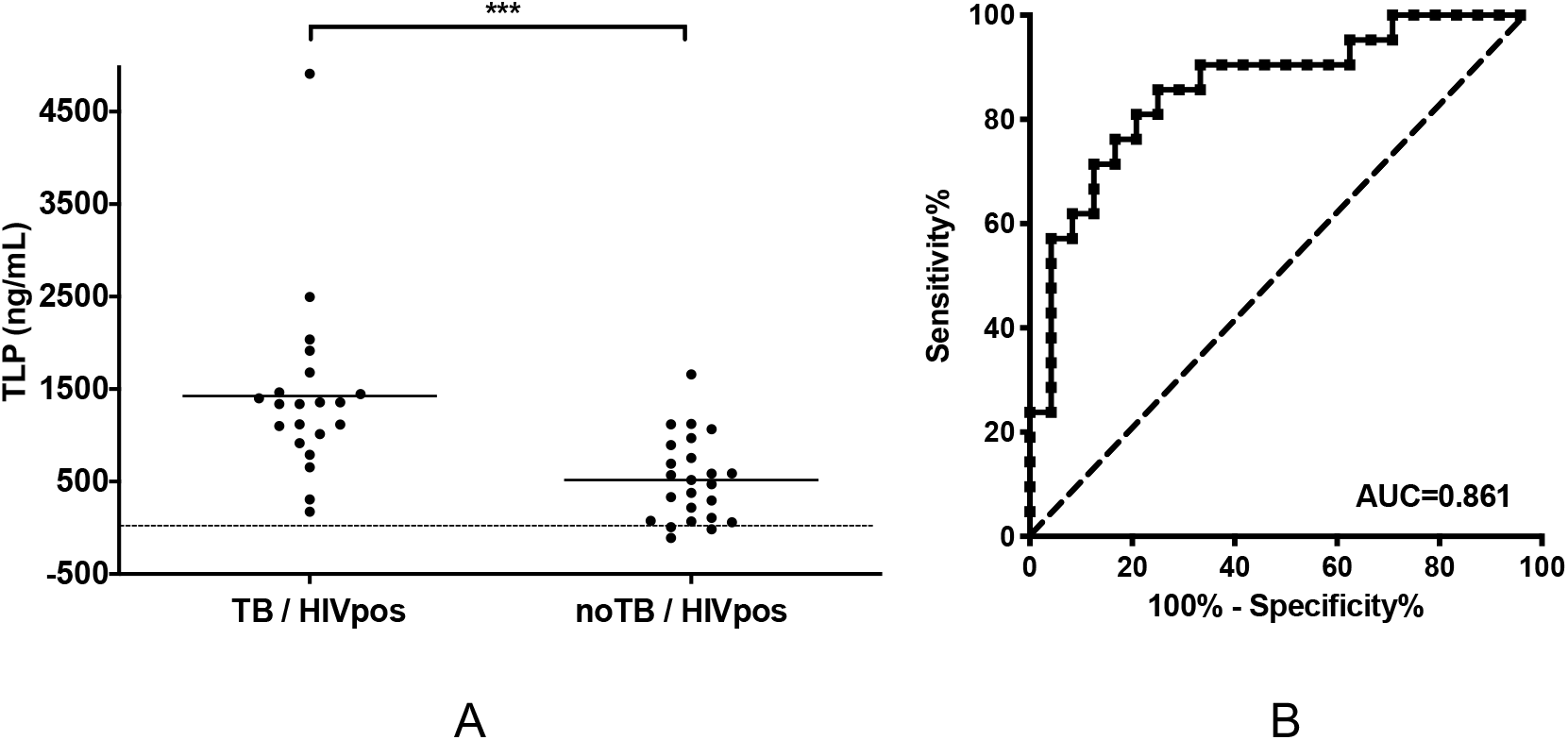
Retrospective analysis of P1041 HIV positive plasma samples. (A) Plasma levels of TLP in HIV positive subjects clinically diagnosed probably having active stage tuberculosis disease (TB) and not having TB (noTB). Individual patients are depicted as dots with group mean. The dashed line represents the detection limit of the assay. *** p<0.001. (B) ROC curve of TLP concentration for active TB versus control subjects of HIV positive patients. AUC, area under curve. The dashed line represents the no-discrimination line from the left bottom to the top right corners.

We also monitored apoB levels, i.e., LDL levels, of all the samples using Human apoB ELISA kits (ab190806, Abcam) and found there was no linear correlation between TLP and apoB levels (Figure S6A), suggesting that plasma LDL fluctuation does not interfere with TLP detection. There is a weak correlation between TLP levels and time before or after diagnosis (Figure S6B). The samples collected before diagnosis may represent disease progression from no infection or infection but without clinical signs of disease to outward manifestation of disease. Samples collected after diagnosis may represent a mixed outcome of disease progression and anti-tubercular treatment. We do not have additional sample sets to demonstrate if anti-tubercular treatment affects TLP levels. However, the discrimination of TLP levels between control subjects and active TB subjects (post diagnosis with treatment less than 40 weeks) was improved with the exclusion of samples collected after 40 weeks of treatment (p<0.0001), which may represent patients clear of TB (Figure S6). ROC (Receiver operating characteristic) analysis provided an AUC (area under the ROC curve) of 0.88 (95% CI=0.76-0.99, p<0.0001). An optimal cutoff of 632 ng/mL gave 67% sensitivity and 93% specificity. The results provide further evidence that TLP levels are associated with active TB disease. Overall, the results clearly highlight the potential of TLP as a biomarker in the diagnosis of active TB in HIV positive children.

## DISCUSSION AND CONCLUSIONS

The wide spectrum of disease observed in children, and the non-specific signs and symptoms especially in young and HIV+ children, contribute to diagnostic delay and missed opportunities to detect TB, which in turn, favors TB disease progression and poor treatment outcomes. There is urgent need to search for a “Rapid Non-sputum-based Biomarker Test for Tuberculosis Detection” [12]. Most of the biomarkers currently studied are host-derived biomarkers including metabolites and cytokines,[31, 32] the levels of which can be affected by many confounding factors. On the other hand, *Mtb* products can be detected directly in blood, sputum or urine, and are increasingly being used for diagnosis.^[33]^ We have characterized a unique biomolecule, *Mtb* modified lipoprotein, which results from a combination of host and *Mtb* pathogen activity. Functionally, TLP increases the flux of lipid into activated macrophages, thereby stimulating their conversion into foamy multi-nucleated macrophages. The TLP-stimulated macrophages accumulate lipid bodies but have a distinct phenotype from MDA-LDL-stimulated macrophages (Figure 2). We reason that detection of this biomolecule in patient plasma may withstand the heterogeneity associated with variations derived from host and be pathogen or disease specific. Thus, TLP detection may be used as TB diagnosis in young children.

The lipoprotein modification can take place in macrophages in contact with *M. tuberculosis*. There are also reports demonstrating that extracellular vesicles carrying bacterial components are released from infected macrophages and circulated beyond the site of infection [34], where host lipoproteins could be modified via contact with the extracellular vesicles. In addition, the intracellular bacilli are able to avoid killing by escaping phagosome-lysosome fusion. In certain individuals, especially children below 5 years of age and immunosuppressed subjects, the extracellular bacilli may disperse to distant metastatic sites via lymphatics and the bloodstream [35], where host lipoproteins may be modified by direct contact with *M. tuberculosis*. Our data demonstrate that the modified lipoprotein in the present study is distinct from other commonly recognized disease associated LDLs, which are typically negatively charged and smaller in particle size compared to native LDL [36].

The present study established a rapid, non-invasive, and robust sandwich ELISA assay for the detection of a novel biomolecule TLP in plasma. We demonstrate that plasma levels of TLP are significantly distinguishable in HIV positive subjects with active TB disease compared to no TB controls (Figure 3). The significant elevation of plasma levels of TLP in HIV positive subjects with active TB disease suggests the detection of TLP associate with TB active disease in HIV positive subjects. The AUC value of 0.86 evidenced the diagnostic value of TLP for TB detection in child. The target product profiles (TPP) adopted by the World Health Organization (WHO) in 2015 for a “Rapid Non-sputum-based Biomarker Test for Tuberculosis Detection” is a diagnostic sensitivity ≥ 66% for microbiologically confirmed pediatric TB and 98% specificity for a pediatric test [12]. The TPP proposed sensitivity is similar to the sensitivity of the Xpert MTB/RIF assay in microbiologically confirmed samples. Our assay renders 57% sensitivity and 96% specificity, which has the potential to meet the WHO TPP for detection of active TB in children, given the highly heterogenous population of P1041 samples, with respect to the bacteriological and clinical status.

Samples used in our study were obtained from IMPAACT P1041 clinical trial. The accuracy of TB diagnosis for the P1041 trial samples is not known due to the absence of good pediatric TB diagnostics. In addition, microbiological confirmation of TB disease was not obtained for any of these patients in the P1041 trial. TB disease subjects used in the present study are patients infected with TB presenting clinical symptoms of disease. The ELISA assays were performed retrospectively and the variance in storage conditions might affect the antigen integrity and therefore the accuracy of the assay. We have insufficient samples to include other clinical characteristics of the patients into data analysis. The clinical characteristics include but not limited to BCG administration, immunosuppression status like CD4 counts, or clinical manifestation of tuberculosis (pulmonary and extra-pulmonary TB). The consideration of those characteristics may help define the limitation of our TLP assay or render improved accuracy with the combination of other clinical factors.

We focused on HIV positive subjects in the current study. TB diagnosis is particularly difficult among HIV co-infected individuals who may have atypical, nonspecific clinical presentation, high rates of smear negative disease and high rates of extrapulmonary TB [37]. P1041 samples were obtained from TB endemic settings, therefore patients were typically being diagnosed at more advanced stages than those living in the US. Therefore, the heterogenous stages of TB disease may contribute to the wide dispersity of TLP levels in TB disease subjects (Figure 3A), which is common among TB diagnostic assays. In addition, the duration of anti-tubercular treatment was not consistent among patients in the present study. Change of LDL modification is a dynamic process and may have the potential to monitor treatment outcome. Although not statistically significant, the slight decrease in the TLP level post diagnosis/ anti-tubercular treatment of 40 weeks may be related to a reduction in microbial load (Figures S6A and S6B). Further studies are required to elucidate the effect of different disease status and/or treatment status on TLP levels in plasma, and to extend our preliminary results to other populations such as HIV negative subjects and adults.

It is a growing notion in the field that a single biomarker will not be sufficient for distinguishing TB status in different patient groups, and multiple biomarkers may be used to increase sensitivity and specificity. Further gains in clinical sensitivity and specificity may be obtained by combining the current assay with other biomarkers or diagnostic tests.

## Supporting information

SupportingInformation

## Data Availability

The datasets used and/or analysed during the current study are available from the corresponding author on reasonable request.

## LIST OF ABBREVIATIONS

TB: tuberculosis
*Mtb*: *Mycobacterium tuberculosis*
ELISA: enzyme-linked immunosorbent assay
FM: foamy macrophages
MDA-LDL: malondialdehyde-modified low density lipoprotein
TLP: *Mycobacterium tuberculosis* modified lipoprotein
ROC: receiver operating characteristic
AUC: area under the ROC curve.

## DECLARATIONS

### Ethics approval and consent to participate

The NIH-IMPAACT Scientific Leadership Group (SLG) reviewed and approved the use of P1041 trial samples under NWCS127. The Wits Health Consortium, University of Witwatersrand, South Africa provided deidentified samples to Stony Brook University for which informed consent had been obtained from a parent and/or legal guardian. The Stony Brook University CORIHS determined the experiments in this manuscript were not human subjects research under either the Common Rule or FDA Regulations.

### Consent for publication

All authors have approved the manuscript for submission.

### Authors’ contributions

SN, MFW, XY, NSS conceived the experiments

SN, MFW, obtained sample resource

XY performed the experiments

XY, NSS wrote the main manuscript text

XY prepared all figures

All authors reviewed the manuscript

## Acknowledgements

We thank Guannan Chen for help with initial LDL purification experiments.

## REFERENCES

1. WHO. 2020. Global tuberculosis report https://apps.who.int/iris/bitstream/handle/10665/336069/9789240013131-eng.pdf Accessed 16 March 2021.

2. Cotton MF, Schaaf HS, Hesseling AC, Madhi SA: HIV and childhood tuberculosis: the way forward. The international journal of tuberculosis and lung disease: the official journal of the International Union against Tuberculosis and Lung Disease 2004, 8(5):675–682.

3. Marais BJ, Gie RP, Obihara CC, Hesseling AC, Schaaf HS, Beyers N: Well defined symptoms are of value in the diagnosis of childhood pulmonary tuberculosis. Archives of disease in childhood 2005, 90(11):1162–1165.

4. Moore DP, Klugman KP, Madhi SA: Role of Streptococcus pneumoniae in hospitalization for acute community-acquired pneumonia associated with culture-confirmed Mycobacterium tuberculosis in children: a pneumococcal conjugate vaccine probe study. The Pediatric infectious disease journal 2010, 29(12):1099–1004.

5. Zar HJ, Tannenbaum E, Apolles P, Roux P, Hanslo D, Hussey G: Sputum induction for the diagnosis of pulmonary tuberculosis in infants and young children in an urban setting in South Africa. Archives of disease in childhood 2000, 82(4):305–308.

6. Walters E, Cotton MF, Rabie H, Schaaf HS, Walters LO, Marais BJ: Clinical presentation and outcome of tuberculosis in human immunodeficiency virus infected children on antiretroviral therapy. BMC pediatrics 2008, 8:1.

7. Jeena PM, Mitha T, Bamber S, Wesley A, Coutsoudis A, Coovadia HM: Effects of the human immunodeficiency virus on tuberculosis in children. Tubercle and lung disease: the official journal of the International Union against Tuberculosis and Lung Disease 1996, 77(5):437–443.

8. Schaaf HS, Gie RP, Beyers N, Smuts N, Donald PR: Tuberculosis in infants less than 3 months of age. Archives of disease in childhood 1993, 69(3):371–374.

9. Jeena PM, Pillay P, Pillay T, Coovadia HM: Impact of HIV-1 co-infection on presentation and hospital-related mortality in children with culture proven pulmonary tuberculosis in Durban, South Africa. The international journal of tuberculosis and lung disease: the official journal of the International Union against Tuberculosis and Lung Disease 2002, 6(8):672–678.

10. Madhi SA, Huebner RE, Doedens L, Aduc T, Wesley D, Cooper PA: HIV-1 co-infection in children hospitalised with tuberculosis in South Africa. The international journal of tuberculosis and lung disease: the official journal of the International Union against Tuberculosis and Lung Disease 2000, 4(5):448–454.

11. Marais BJ, Gie RP, Schaaf HS, Hesseling AC, Obihara CC, Starke JJ, Enarson DA, Donald PR, Beyers N: The natural history of childhood intra-thoracic tuberculosis: a critical review of literature from the pre-chemotherapy era. The international journal of tuberculosis and lung disease: the official journal of the International Union against Tuberculosis and Lung Disease 2004, 8(4):392–402.

12. Denkinger CM, Kik SV, Cirillo DM, Casenghi M, Shinnick T, Weyer K, Gilpin C, Boehme CC, Schito M, Kimerling M et al: Defining the needs for next generation assays for tuberculosis. J Infect Dis 2015, 211 Suppl 2:S29–38.

13. Abdalla AE, Ejaz H, Mahjoob MO, Alameen AAM, Abosalif KOA, Elamir MYM, Mousa MA: Intelligent Mechanisms of Macrophage Apoptosis Subversion by Mycobacterium. Pathogens 2020, 9(3).

14. Genoula M, Marin Franco JL, Dupont M, Kviatcovsky D, Milillo A, Schierloh P, Morana EJ, Poggi S, Palmero D, Mata-Espinosa D et al: Formation of Foamy Macrophages by Tuberculous Pleural Effusions Is Triggered by the Interleukin-10/Signal Transducer and Activator of Transcription 3 Axis through ACAT Upregulation. Front Immunol 2018, 9:459.

15. Russell DG, Cardona PJ, Kim MJ, Allain S, Altare F: Foamy macrophages and the progression of the human tuberculosis granuloma. Nat Immunol 2009, 10(9):943–948.

16. Viljoen A, Blaise M, de Chastellier C, Kremer L: MAB_3551c encodes the primary triacylglycerol synthase involved in lipid accumulation in Mycobacterium abscessus. Mol Microbiol 2016, 102(4):611–627.

17. Caire-Brandli I, Papadopoulos A, Malaga W, Marais D, Canaan S, Thilo L, de Chastellier C: Reversible lipid accumulation and associated division arrest of Mycobacterium avium in lipoprotein-induced foamy macrophages may resemble key events during latency and reactivation of tuberculosis. Infect Immun 2014, 82(2):476–490.

18. Guerrini V, Gennaro ML: Foam Cells: One Size Doesn’t Fit All. Trends Immunol 2019, 40(12):1163–1179.

19. Agarwal P, Combes TW, Shojaee-Moradie F, Fielding B, Gordon S, Mizrahi V, Martinez FO: Foam Cells Control Mycobacterium tuberculosis Infection. Front Microbiol 2020, 11:1394.

20. Watanabe K, Nakazato Y, Saiki R, Igarashi K, Kitada M, Ishii I: Acrolein-conjugated low-density lipoprotein induces macrophage foam cell formation. Atherosclerosis 2013, 227(1):51–57.

21. Holvoet P, Vanhaecke J, Janssens S, Van de Werf F, Collen D: Oxidized LDL and malondialdehyde-modified LDL in patients with acute coronary syndromes and stable coronary artery disease. Circulation 1998, 98(15):1487–1494.

22. Knappik A, Ge L, Honegger A, Pack P, Fischer M, Wellnhofer G, Hoess A, Wolle J, Pluckthun A, Virnekas B: Fully synthetic human combinatorial antibody libraries (HuCAL) based on modular consensus frameworks and CDRs randomized with trinucleotides. J Mol Biol 2000, 296(1):57–86.

23. Prassler J, Thiel S, Pracht C, Polzer A, Peters S, Bauer M, Norenberg S, Stark Y, Kolln J, Popp A et al: HuCAL PLATINUM, a synthetic Fab library optimized for sequence diversity and superior performance in mammalian expression systems. J Mol Biol 2011, 413(1):261–278.

24. Madhi SA, Nachman S, Violari A, Kim S, Cotton MF, Bobat R, Jean-Philippe P, McSherry G, Mitchell C, Team PS: Primary isoniazid prophylaxis against tuberculosis in HIV-exposed children. The New England journal of medicine 2011, 365(1):21–31.

25. A randomized, double blind, placebo controlled trial to determine the efficacy of isoniazid (INH) in preventing tuberculosis disease and latent tuberculosis infection among infants with perinatal exposure to HIV. [https://impaactnetwork.org/DocFiles/P1041/P1041V2_11Jul07.pdf]

26. Gupta A, Montepiedra G, Gupte A, Zeldow B, Jubulis J, Detrick B, Violari A, Madhi S, Bobat R, Cotton M et al: Low Vitamin-D Levels Combined with PKP3-SIGIRR-TMEM16J Host Variants Is Associated with Tuberculosis and Death in HIV-Infected and -Exposed Infants. PloS one 2016, 11(2):e0148649.

27. Yoshida M, Higashi K, Kobayashi E, Saeki N, Wakui K, Kusaka T, Takizawa H, Kashiwado K, Suzuki N, Fukuda K et al: Correlation between images of silent brain infarction, carotid atherosclerosis and white matter hyperintensity, and plasma levels of acrolein, IL-6 and CRP. Atherosclerosis 2010, 211(2):475–479.

28. Puissegur MP, Lay G, Gilleron M, Botella L, Nigou J, Marrakchi H, Mari B, Duteyrat JL, Guerardel Y, Kremer L et al: Mycobacterial lipomannan induces granuloma macrophage fusion via a TLR2-dependent, ADAM9-and beta1 integrin-mediated pathway. J Immunol 2007, 178(5):3161–3169.

29. Nezami N, Ghorbanihaghjo A, Rashtchizadeh N, Argani H, Tafrishinejad A, Ghorashi S, Hajhosseini B: Atherogenic changes of low-density lipoprotein susceptibility to oxidation, and antioxidant enzymes in pulmonary tuberculosis. Atherosclerosis 2011, 217(1):268–273.

30. Sacks FM, Moye LA, Davis BR, Cole TG, Rouleau JL, Nash DT, Pfeffer MA, Braunwald E: Relationship between plasma LDL concentrations during treatment with pravastatin and recurrent coronary events in the cholesterol and recurrent events trial. Circulation 1998, 97(15):1446–1452.

31. Weiner J, Kaufmann SH: Recent advances towards tuberculosis control: vaccines and biomarkers. Journal of internal medicine 2014.

32. Walzl G, Ronacher K, Hanekom W, Scriba TJ, Zumla A: Immunological biomarkers of tuberculosis. Nature reviews Immunology 2011, 11(5):343–354.

33. Goletti D, Lee MR, Wang JY, Walter N, Ottenhoff THM: Update on tuberculosis biomarkers: from correlates of risk, to correlates of active disease and of cure from disease. Respirology 2018, 23(5):455–466.

34. Athman JJ, Wang Y, McDonald DJ, Boom WH, Harding CV, Wearsch PA: Bacterial membrane vesicles mediate the release of Mycobacterium tuberculosis lipoglycans and lipoproteins from infected macrophages. J Immunol 2015, 195(3):1044–1053.

35. Grosset J: Mycobacterium tuberculosis in the extracellular compartment: an underestimated adversary. Antimicrob Agents Chemother 2003, 47(3):833–836.

36. Orekhov AN: LDL and foam cell formation as the basis of atherogenesis. Curr Opin Lipidol 2018, 29(4):279–284.

37. Scott L, da Silva P, Boehme CC, Stevens W, Gilpin CM: Diagnosis of opportunistic infections: HIV co-infections - tuberculosis. Curr Opin HIV AIDS 2017, 12(2):129–138.

