## SupportingInformation for "Retrospective feasibility study of *Mycobacterium tuberculosis* modified lipoprotein as a potential biomarker for TB detection in children"

Running title: Retrospective Mtb lipoprotein biomarker in children

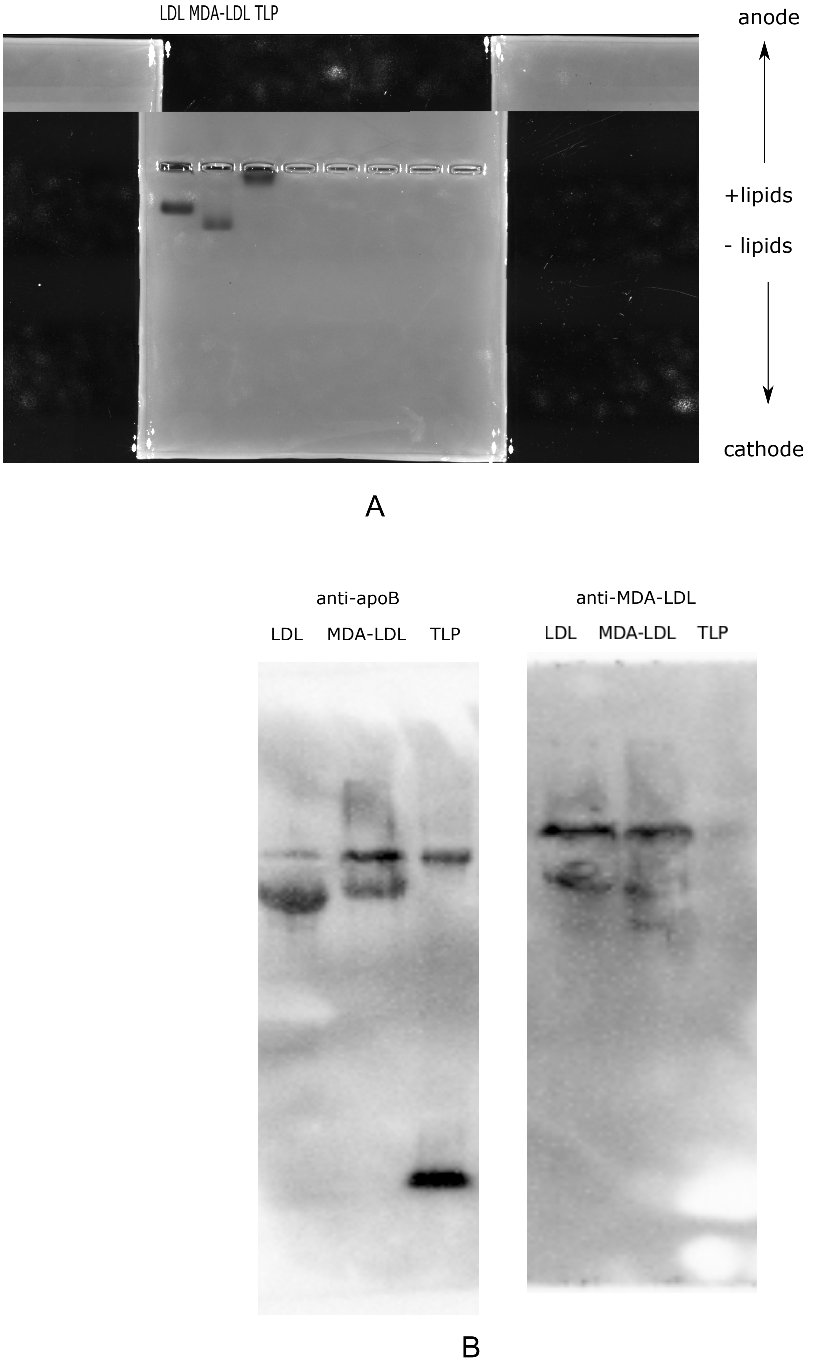


**Figure S1.** Full-length blots for Figure 1**.** (A) an agarose gel electrophoresis showing TLP is larger in size and more positively charged than native LDL. The top of the fragile gel cracked and moved during scanning (B) Immunoblots showing TLP is recognized by anti- apolipoprotein B (apoB) antibodies (left) but not anti- MDA-LDL antibodies (right).

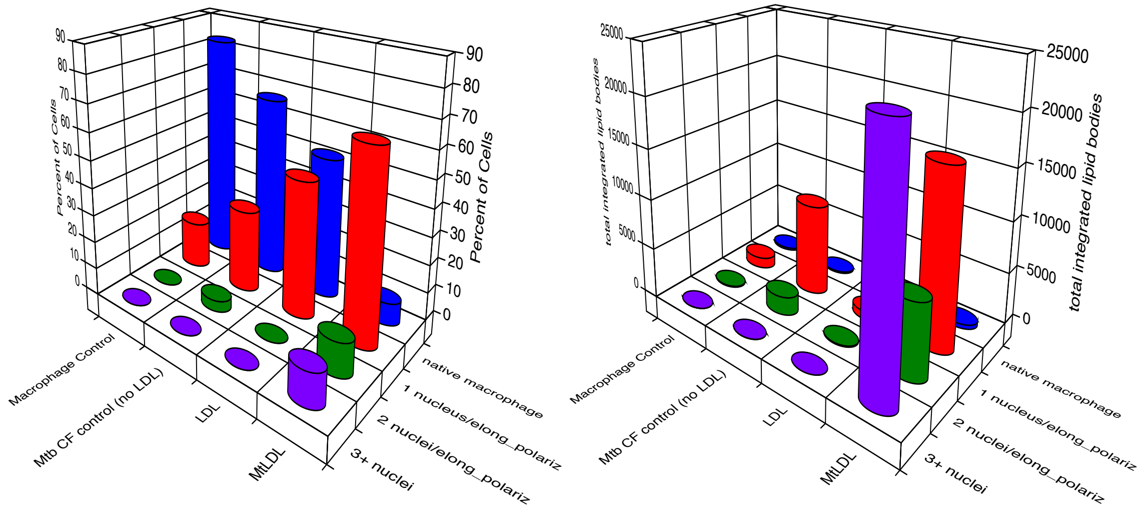


**Figure S2. TLP induces lipid body accumulation in macrophages.** (A) Graph depicting higher level of formation of multi-nucleated THP-1 macrophages by TLP in comparison to LDL. (B) Graph depicting formation of higher concentration of lipid bodies in THP-1 macrophages; by TLP in comparison to LDL.

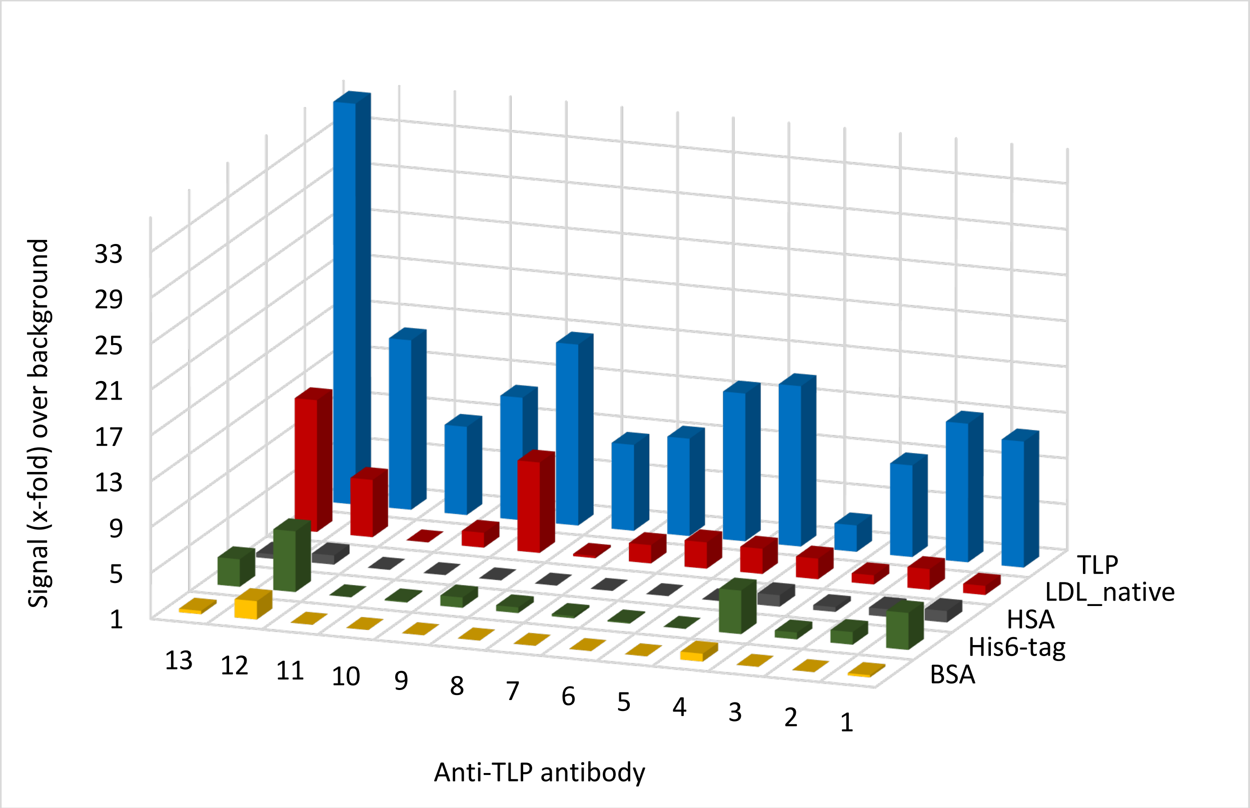


**Figure S3.** illustrates a method using indirect ELISA for determining specificity of 13 selected TLP binding Fab bivalent fragments that were identified from HuCAL phage display library screen; wherein, 8 Fab bivalent fragments showed high signal/background ratio as depicted on y-axis as shown higher signal in TLP coated wells as compared to native LDL, HSA, His6-tag and BSA.

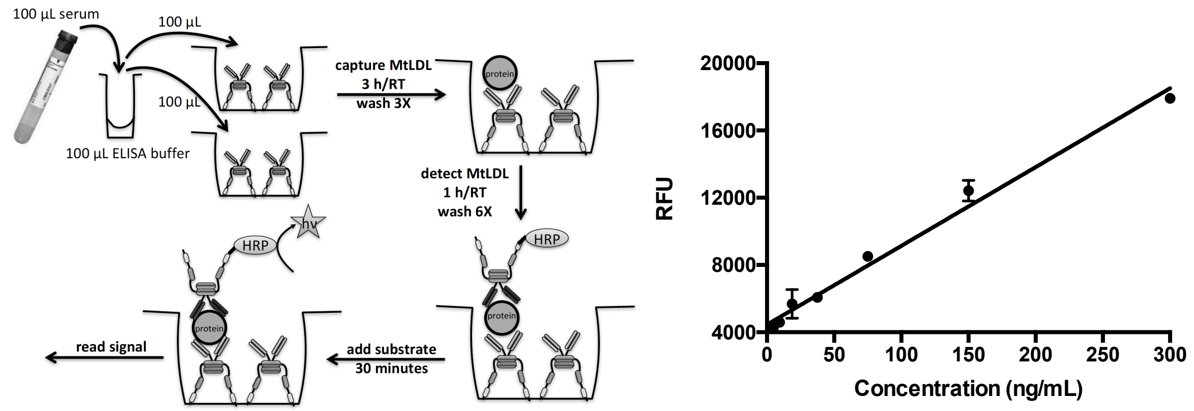


A B

**Figure S4.** (A) Example schematic of a sandwich ELISA to detect TLP levels in 100 μL infant serum samples from diagnosis of M. tuberculosis infection, using sandwich pair of Fab fragments 8 and 6. (B) Calibration curve of sandwich ELISA method showing linear correlation between antigen specific binding of a pairwise combination of TLP specific Fab fragments 6 and 8 (y-axis) to increasing concentration of TLP in a TLP spiked cord blood serum sample (x-axis). A lower detection limit of 4-5 ng/mL in serum is shown.

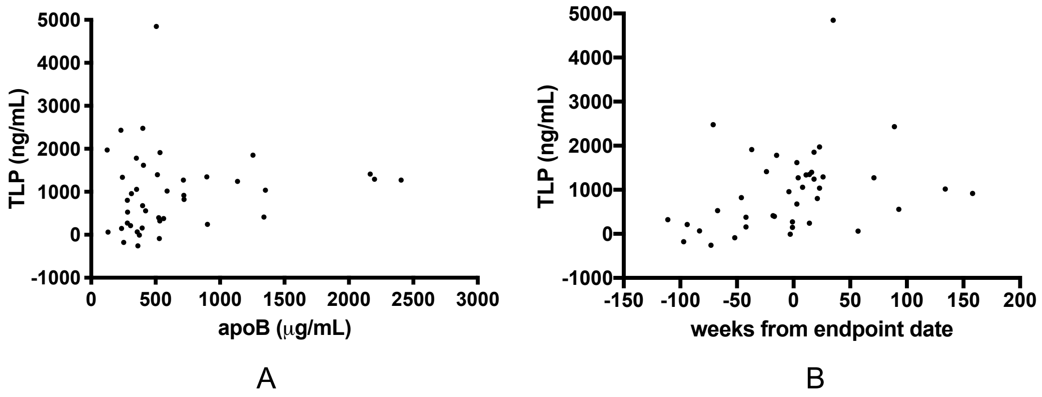


**Figure S5.** Plasma levels of TLP versus apoB levels (A) and time (in weeks) pre- or post-diagnosis (B). (A) Person r=0.131 and p=0.413. (B) Person r=0.310 and p=0.048.

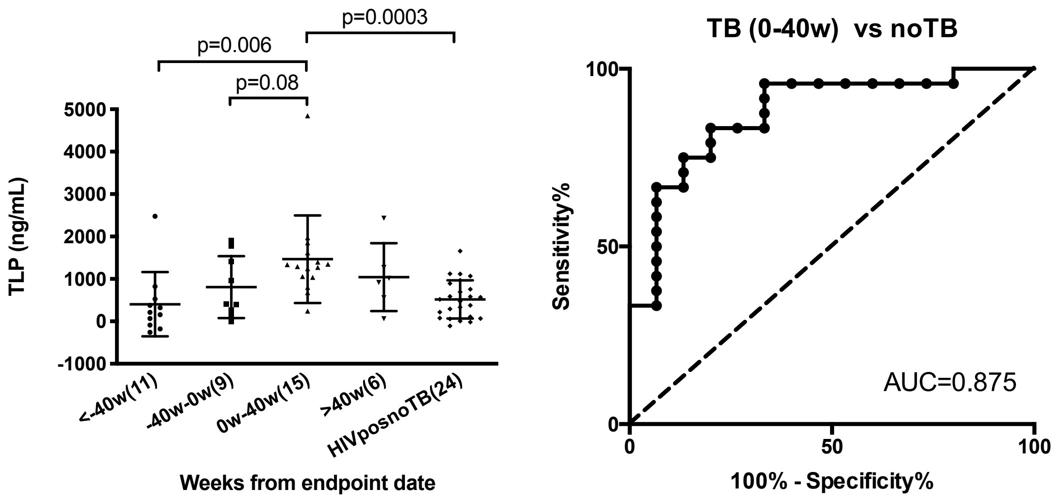


A B

**Figure S6.** Retrospective analysis of P1041 HIVpos plasma samples. (A) Longitudinal presentation (weeks before < 0w or after diagnosis >0w) of plasma levels of TLP in HIV positive subjects, with respect to noTB control subjects. Individual patients are depicted as dots with group mean. Error bars are the standard error of the mean. (B) ROC curve of TLP concentration for plasma samples collected 0-40 weeks after diagnosis versus control subjects, HIV positive. AUC, area under curve. The dash line represents the no-discrimination line from the left bottom to the top right corners.

**Table S1**. Dynamic light scattering analysis of LDL, MDA-LDL and TLP

| Sample | Effective Diameter (nm) | Polydispersity |
|---|---|---|
| LDL | 25.3 ± 0.2 | 0.215 ± 0.005 |
| MDA-LDL | 57.0 ± 0.4 | 0.218 ± 0.006 |
| TLP | 61.0 ± 1.4 | 0.337 ± 0.002 |

**Table S2.** Amino acid sequences of AbD28582 and AbD28580.

Underlined sequences represent complementarity determining regions (CDR). Constant domains CH1 and CL sequence are in italics, dimerization domain sequence (AP) is double underlined, linker sequences are in bold, FLAG® tag is underlined with dashed line and His6 tag is in underlined with thick line.

| Amino acid sequence of Fd chain and tags, AbD28580 |
|---|
| EVQLLESGGGLVQPGGSLRLSCAASGFTFRGYYMSWVRQAPGKGLEWVSSISGFSSNTYY<br>ADSVKGRFTISRDNSKNTLYLQMNSLRAEDTAVYYCARVRYLAYAFDYWGQGTLVTVSS*AS*<br>*TKGPSVFPLAPSSKSTSGGTAALGCLVKDYFPEPVTVSWNSGALTSGVHTFPAVLQSSGLY*<br>*SLSSVVTVPSSSLGTQTYICNVNHKPSNTKVDKKVEPKS*EFKAEMPVLENRAAQGDITTPGG<br>ARRLTGDQTAALRDSLSDKPAKNIILLIGDGMGDSEITAARNYAEGAGGFFKGIDALPLTGQY<br>THYALNRKTGKPDYVTSSAASATAWSTGVKTYNGALGVDIHEKDHPTILEMAKAAGLATGNV<br>STAELQDATPAALVAHVTSRKCYGPSATSEKCPGNALEKGGKGSITEQLLNARADVTLGGG<br>AKTFAETATAGEWQGKTLREQAQARGYQLVSDAASLNSVTEANQQKPLLGLFADGNMPVR<br>WLGPKATYHGNIDKPAVTCTPNPQRNDSVPTLAQMTDKAIELLSKNEKGFFLQVEGASIDKQ<br>DHAANPCGQIGETVDLDEAVQRALEFAKKEGNTLVIVTADHAHASQIVAPDTKAPGLTQALNT<br>KDGAVMVMSYGNSEEDSQEHTGSQLRIAAYGPHAANVVGLTDQTDLFYTMKAALGLKGAP<br>DYKDDDDKGAPHHHHHH |
| Amino acid sequence of light chain, AbD28580 |
| DIELTQPPSVSVSPGQTASITCSGDSLPDKRAYWYQQKPGQAPVLVIYGDSHRPSGIPERFS<br>GSNSGNTATLTISGTQAEDEADYYCSSWGSRTWVFGGGTKLTVLGQ*PKAAPSVTLFPPSSE*<br>*ELQANKATLVCLISDFYPGAVTVAWKADSSPVKAGVETTTPSKQSNNKYAASSYLSLTPEQW*<br>*KSHRSYSCQVTHEGSTVEKTVAPTEA* |
| Amino acid sequence of Fd chain and tags, AbD28582 |
| QVQLVQSGAEVKKPGSSVKVSCKASGGTFSGYYISWVRQAPGQGLEWMGGIIPISGRANYA<br>QKFQGRVTITADESTSTAYMELSSLRSEDTAVYYCARSRSYYHFDLWGQGTLVTVSS*ASTK*<br>*GPSVFPLAPSSKSTSGGTAALGCLVKDYFPEPVTVSWNSGALTSGVHTFPAVLQSSGLYSLS*<br>*SVVTVPSSSLGTQTYICNVNHKPSNTKVDKKVEPKS*EFKAEMPVLENRAAQGDITTPGGARR<br>LTGDQTAALRDSLSDKPAKNIILLIGDGMGDSEITAARNYAEGAGGFFKGIDALPLTGQYTHY<br>ALNRKTGKPDYVTSSAASATAWSTGVKTYNGALGVDIHEKDHPTILEMAKAAGLATGNVSTA<br>ELQDATPAALVAHVTSRKCYGPSATSEKCPGNALEKGGKGSITEQLLNARADVTLGGGAKTF<br>AETATAGEWQGKTLREQAQARGYQLVSDAASLNSVTEANQQKPLLGLFADGNMPVRWLGP<br>KATYHGNIDKPAVTCTPNPQRNDSVPTLAQMTDKAIELLSKNEKGFFLQVEGASIDKQDHAA |

| |
|---|
| NPCGQIGETVDLDEAVQRALEFAKKEGNTLVIVTADHAHASQIVAPDTKAPGLTQALNTKDGAVMVMSYGNSEEDSQEHTGSQLRIAAYGPHAANVVGLTDQTDLFYTMKAALGLKGAPDYKDDDDKGAPHHHHHH |
| Amino acid sequence of light chain, AbD28582 |
| DIALTQPASVSGSPGQSITISCTGTSSDVGRYNSVSWYQQHPGKAPKLMIYRVSKRPSGVSNRFSGSKSGNTASLTISGLQAEDEADYYCQSWASLSNVVFGGGTKLTVLGQ*PKAAPSVTLFPPSSEELQANKATLVCLISDFYPGAVTVAWKADSSPVKAGVETTTPSKQSNNKYAASSYLSLTPEQWKSHRSYSCQVTHEGSTVEKTVAPTEA* |
